# Development and Evaluation of A CRISPR-based Diagnostic For 2019-novel Coronavirus

**DOI:** 10.1101/2020.02.22.20025460

**Authors:** Tieying Hou, Weiqi Zeng, Minling Yang, Wenjing Chen, Lili Ren, Jingwen Ai, Ji Wu, Yalong Liao, Xuejing Gou, Yongjun Li, Xiaorui Wang, Hang Su, Bing Gu, Jianwei Wang, Teng Xu

## Abstract

**Background:** The recent outbreak of infections by the 2019 novel coronavirus (2019-nCoV), the third zoonotic CoV has raised great public health concern. The demand for rapid and accurate diagnosis of this novel pathogen brought significant clinical and technological challenges. Currently, metagenomic next-generation sequencing (mNGS) and reverse-transcription PCR (RT-PCR) are the most widely used molecular diagnostics for 2019-nCoV.

**Methods:** 2019-nCoV infections were confirmed in 52 specimens by mNGS. Genomic information was analyzed and used for the design and development of an isothermal, CRISPR-based diagnostic for the novel virus. The diagnostic performance of CRISPR-nCoV was assessed and also compared across three technology platforms (mNGS, RT-PCR and CRISPR)

**Results:** 2019-nCoVs sequenced in our study were conserved with the Wuhan strain, and shared certain genetic similarity with *SARS-CoV*. A high degree of variation in the level of viral RNA was observed in clinical specimens. CRISPR-nCoV demonstrated a near single-copy sensitivity and great clinical sensitivity with a shorter turn-around time than RT-PCR.

**Conclusion:** CRISPR-nCoV presents as a promising diagnostic option for the emerging pathogen.

## INTRODUCTION

Since the beginning of 2020, a surging number of pneumonia caused by infections of a novel coronavirus (2019-nCoV) have been identified in China, especially Wuhan, a metropolitan city with a population of over 10 million.[1, 2] In recent days, the outbreak has affected multiple countries and caused worldwide impact.[3-6] Currently, nucleic-acid based tests have been widely used as the reference method for the diagnosis of 2019-nCoV.[7] As of now, more than 30,000 individuals have been identified with 2019-nCoV infection. As the epidemic develops, there are increasing demands for rapid and sensitive diagnostics for the novel pathogen.

Coronaviruses (CoVs) are positive-sense, single-strand RNA viruses, with four major structural proteins including spike (S), membrane (M), envelop (E) and nucleoprotein (N).[8, 9] Prior to 2019-nCoV, there were six CoVs that were known to be pathogenic to humans: *HCoV-OC43, HCoV-NL63, HCoV-HKU1, HCoV-229E* and highly transmissible and pathogenic *SARS-CoV* and *MERS-CoV*. [10-12]

The epidemic of 2019-nCoV was first discovered by metagenomic next-generation sequencing (mNGS) in which the novel virus was found to be a new pathogenic member of the betacoronavirus genus but shared only about 79% in genetic similarity with *SARS*.[8, 9] Currently, metagenomics and RT-PCR are two molecular approaches most commonly used diagnostics for this novel virus. [13,14] However, the diagnostic performance of different molecular methods have not been investigated.

Although CRISPR/Cas has been widely used as a programmable tool for gene editing since 2013, the collateral, promiscuous cleavage activities of a unique group of Cas nucleases only discovered recently and harnessed for in vitro nucleic acid detection.[15-19]

Here, to address this question and the expanding clinical needs, we developed CRISPR-nCoV, a rapid assay for 2019-nCoV detection, and compared the diagnostic performance among three different technological platforms: metagenomic sequencing, RT-PCR and CRISPR. To our knowledge, this is the first report on cross-platform comparison and the evaluation of an isothermal, CRISPR-based assay for 2019-nCoV that’s rapid, sensitive and with low instrument requirement.

## MATERIALS AND METHODS

### Study participants and sample collection

This study used excess RNA samples from patients with suspected 2019-nCoV infection based on clinical, chest imaging and epidemiological evidence. No patient identifiable information was collected. The only data collected from the samples were types of specimens (nasopharyngeal swab or bronchoalveolar lavage fluid), the concentrations and volumes of the purified total RNA. A total of sixty-one 2019-nCoV-suspected samples were included in this study. Among which, 52 was confirmed positive by mNGS. This study was approved by the ethical review committee of Institute of Pathogen Biology, Chinese Academy of Medical Sciences & Peking Union Medical College. Written informed consent was waived given the context of emerging infectious diseases.

### mNGS Assay for 2019-nCoV

The RNA concentrations were measured by a Qubit Fluorometer (Thermo Fisher Scientific, Carlsbad, CA, USA). Sequencing libraries were constructed by a transposase-based methodology with ribosomal RNA depletion (Vision Medicals, China). Sequencing was performed on a Nextseq sequencer (Illumina, San Diego, CA). At least 10 million single-end 75bp reads were generated for each sample. Quality control processes included removal of low-complexity reads, low-quality reads and short reads, as well as adapter trimming. Reads derived from host genome were then removed. Taxonomic assignment of the clean reads was performed by aligning against the reference databases, including archaea, bacteria, fungi, human, plasmid, protozoa, univec, and virus sequences. A negative control sample was processed and sequenced in parallel for each sequencing run for contamination control.

### Phylogenetic analysis

Phylogenetic trees were constructed based on the genome sequences by means of the maximum-likelihood method. Alignment of multiple sequences was performed with the ClustalW program (MEGA software, version 7.0.14).

### Cas13a protein and other reagents

The open reading frame (ORF) of Cas13a was synthesized after codon optimization. The Cas13a ORF was then cloned into expression vector Pc013 and transfected into E. coli BL21, which were first grown at 37°C and incubated with IPTG at 16°C. Proteins were purified from lysed bacteria using the Ni-NTA protocol. Aliquots of purified protein were stored at −80°C. Other reagents were purchased from Sangon Co., Ltd. (Shanghai, China), including DTT (A100281), EDTA (A100105), TritonX-100 (A110694), NP-40 (A100109), Chelex-100 (C7901) etc.

### Strains and human DNA

Pure human DNA were purchased from Solarbio Co.,Ltd. (Beijing, China), and eluted in nuclease-free water. *Bacterial and viral* strains were purchased from the American Type Culture Collection (ATCC), China General Microbiological Culture Collection Center (CGMCC) or BDS (Guangzhou, China).

### Oligos and gRNA

Primer with an appended T7 promoter used in the RPA amplification for *Orf1ab* amplification were forward primer 5’-TAAT ACGA CTCA CTAT AGGG ACAT AAAC AAGC TTTG TGAA GAAA TGCT GGAC-3’ and reverse primer 5’-TTGA GCAG TAGC AAAA GCTG CATA TGAT GGAA GG-3’. gRNA for *Orf1ab* (5’-GGGG AUUU AGAC UACC CCAA AAAC GAAG GGGA CUAA AACA AACU CUGA GGCU AUAG CUUG UAAG GUU-3’) and ssRNA probe (5’-6-FAM-UUUU UC-BHQ1) were used for the CRISPR detection following RPA amplification.

### CRISPR-nCoV

The CRISPR-nCoV test combines an Recombinase Polymerase Amplification (RPA) step and a following T7 transcription and Cas13 detection step as described previously. Briefly, reactions containing 2.5µl of sample, 0.4 µM of each primer, 1× reaction buffer, 14 mM of magnesium acetate and the RT-RPA enzyme mix were incubated at 42°C for 30 min. After that, the CRISPR reaction mix consisting of the amplification product, 33.3 nM of gRNA, 66.7nM of Cas13, 5mM of each NTP, 1μl T7 RNA polymerase (New England Biolabs) and 166 nM of ssRNA reporter was incubated at 42°C and monitored for fluorescence signal. Fluorescent signals were collected for the duration of 10 min.

### Plasmid construction

A 420bp genomic fragment of *Orf1ab* encompassing 310bp upstream and 82bp downstream of the gRNA target site was synthesized and inserted into pUC57. These sequences represented 11788-12207bp in the 2019-nCoV genome. This *Orf1ab* plasmid was purfied and used as positive control.

### Evaluation of limit of detection (LoD)

For the evaluation of LoD by the number of DNA copies, DNA of the 2019-nCoV plasmid was purified and the concentration was determined by a Qubit (Thermo Fisher, Massachusetts). The copy number concentration was then calculated based the weight and the length of fragment. Serial dilution with nuclease-free water was done to achieve desired concentrations. 2.5μl of extracted DNA at each titer was used as templates. Ten replicates were performed at each data point near detection limit.

### Statistical analysis

Comparative analysis was conducted by Pearson χ2 test, Fisher exact test, the Student’s t-test or log-rank test where appropriate. Data analyses were performed using SPSS 22.0 software. P values <0.05 were considered significant, and all tests were 2-tailed unless indicated otherwise.

## RESULTS

### Identification of 2019-nCoV by mNGS

In order to develop a targeted assay for the novel virus, we obtained total RNA samples from 61 cases suspected for 2019-nCoV infection and subjected them to metagenomic next-generation sequencing (mNGS), the method by which this novel virus was initially identified. Briefly, RNA was reverse-transcribed into cDNA to prepare for the sequencing library. Each library was subjected to high-throughput sequencing. Sequenced reads were aligned to a curated database for taxonomic classification and identification of 2019-nCoV genomic sequences.

Among the 61 suspected nCoV specimens, we were able to confirm 52 cases with read numbers mapping to the novel virus across 6 orders of magnitudes (median read of 1,484, from 2 to 19,016,501). The median genome coverage and sequencing depth was 46.8% (2.8%-100%) and 12.0× (1.0×-7870.1×), respectively (Figure 1A). These findings indicate a high degree of variation in viral loads of nCoV infections. As shown in the phylogenetic trees, 2019-nCoV genome identified in our specimens were highly conserved with the Wuhan strain and closest to *SARS-CoV* (Figure 1B).

**Figure 1.**
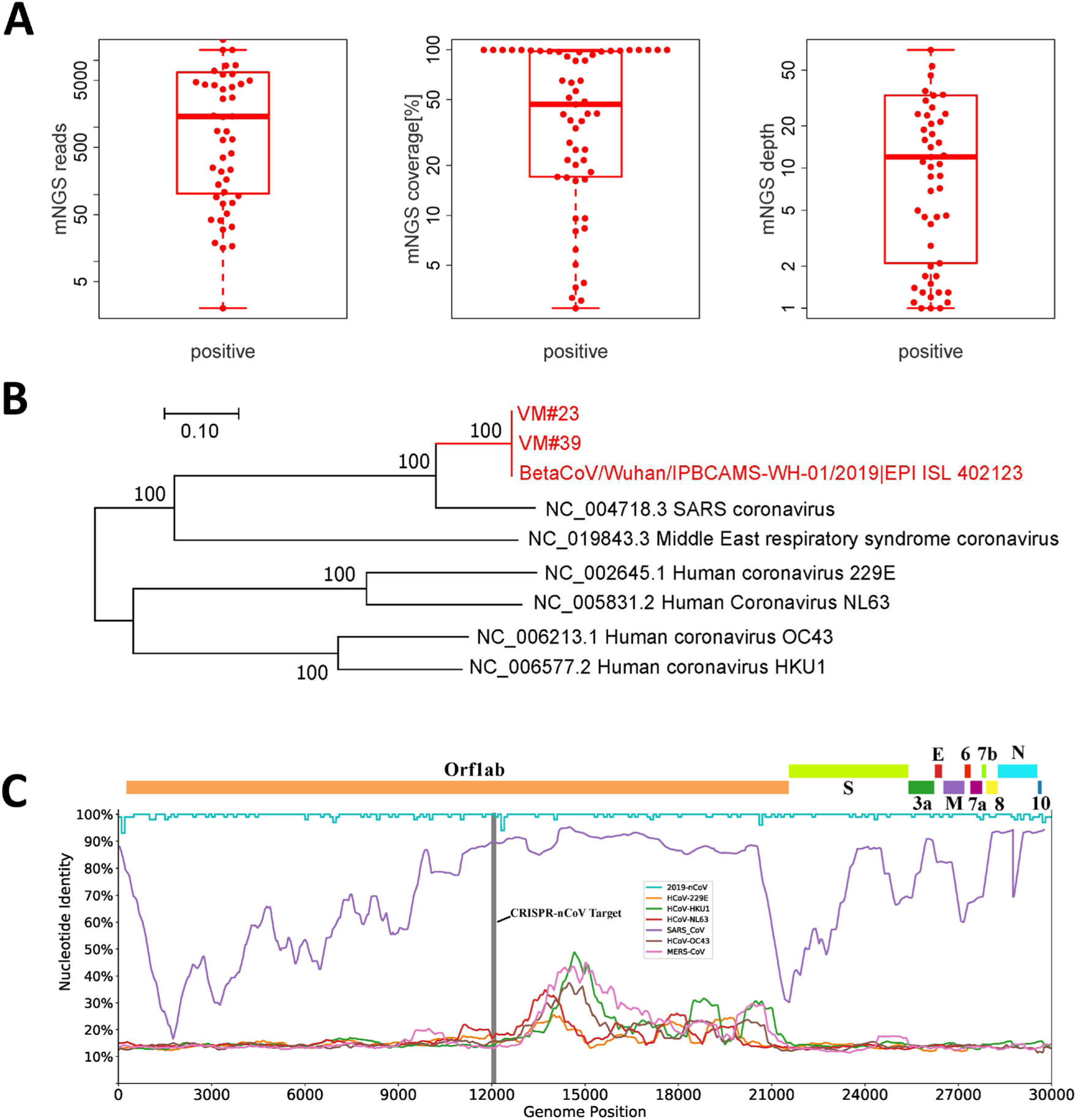
(A) Number of mNGS reads mapped to the genome of 2019-nCoV in 52 positive cases; (B) Phylogenetic tree of 2019-nCoVs and other pathogenic CoVs; (C) 2019-nCoV gene structure and CRISPR target region. Nucleotide identity among 52 cases of 2019-nCoV cases included in this study, between 2019-nCoV (NC_045512.2) and other pathogenic CoVs. Brief gene locations are presented above and the target region for CRISPR-nCoV is indicated in Grey.

With these genomic information, we aimed to identify target regions of 2019-nCoV by searching for sequences that were i) within the *Orf1ab, N* or *E* genes of the viral genome; ii) conserved among strains of the novel virus; iii) differentiable from other pathogenic coronaviruses. By analyzing the genetic similarity among the 52 cases 2019-nCoV and other pathogenic CoVs (Figure 1C), we identified two potential target sequences in *Orf1ab* and one in the *N* gene (data not shown).

### Development of CRISPR-nCoV

We seek to develop a rapid, highly sensitive and simple-to-use assay by taking advantage of both the polymerase-mediated DNA amplification by RPA and the CRISPR/Cas-mediated enzymatic signal amplification for improved sensitivity (Figure 2A). Moreover, the isothermal nature of such an assay abolished the demand for sophiscated instruments such as thermal cyclers as for PCR-based assays.

**Figure 2.**
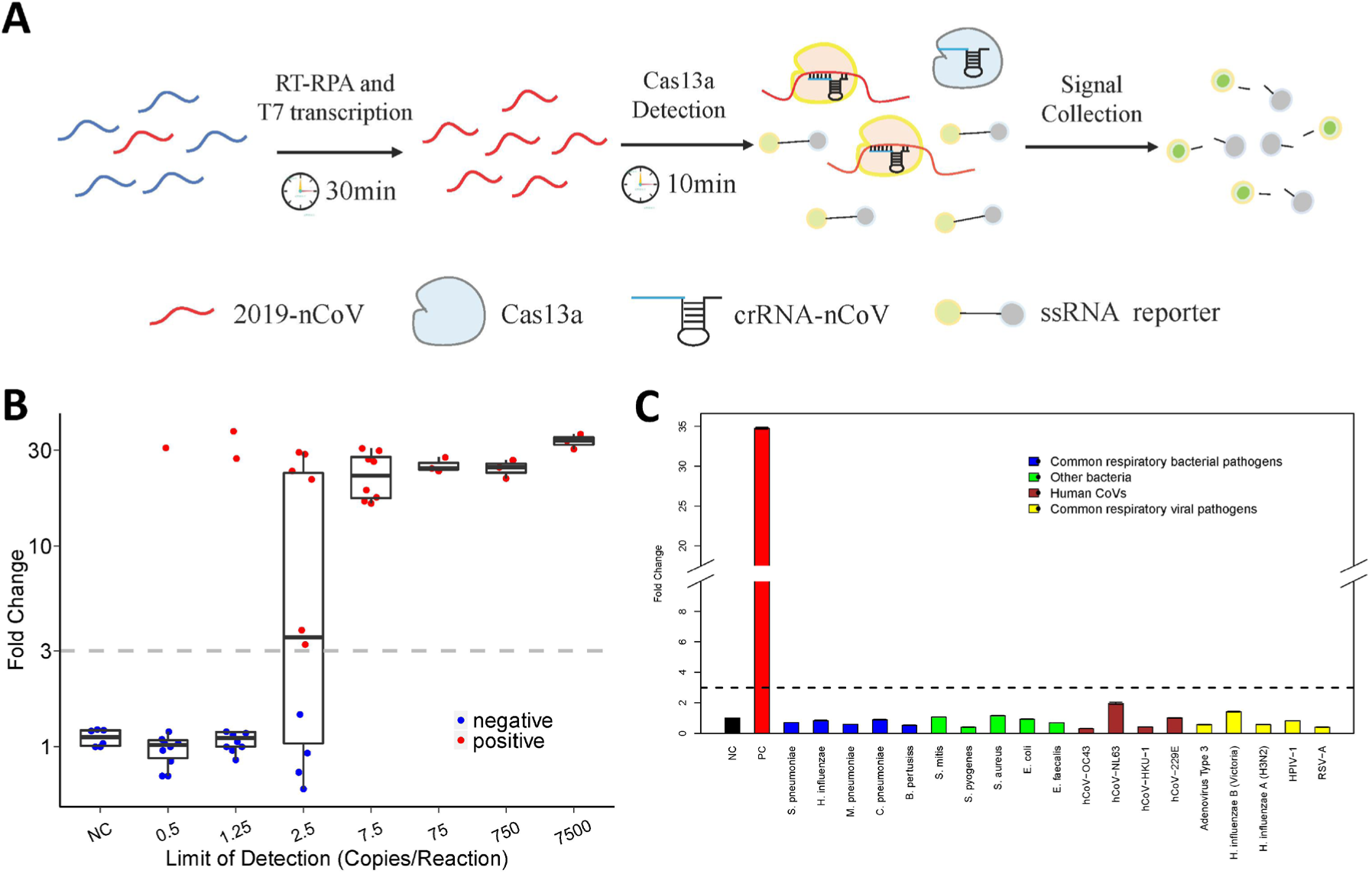
(A) Schematic diagram of CRISPR-nCoV. The collateral nuclease activity of Cas proteins are activated upon specific binding of gRNA to the *Orf1ab* gene. Fluorescent signal produced from cleaved probes is captured and indicates the presence of 2019-nCoV. (B-C) Analytical assessment of the sensitivity and specifcity of CRISPR-nCoV. Evaluation was performed by testing contrived samples with indicated titers of 2019-nCoV (B), and various microbes as interefering materials (C).

Based on the three potential target sequences we identified, multiple sets of RPA primers and CRISPR gRNAs were designed and screened. Among these, the set that targeted *Orf1ab* showed the best overall performance of sensitivity and specificity, and therefore, was used to develop CRISPR-nCoV in this study for further evaluation (data not shown).

We then sought to determine its analytical sensitivity by serial dilution at various concentrations. As shown in Figure 2B, CRISPR-nCoV consistently detected 7.5 copies of 2019-nCoV in all 10 replicates, 2.5 copies in 6 out of 10, and 1.25 copies in 2 out of 10 runs. These data indicate that CRISPR-nCoV had a near single-copy sensitivity. To confirm its specificity, we tested CRISPR-nCoV with DNA from human cells as well as a panel of microbes including i) bacteria commonly found in respiratory infections: *S. pneumonia, H. influenza, M. pneumonia, C. pneumonia, B. pertusiss;* ii) human Coronaviruses: *HCoV-OC43, HCoV-NL63, HCoV-HKU1, HCoV-229E;* iii) other viruses commonly found in respiratory infections: *Adenovirus Type-3, H. Influenza B (Victoria), H influenza A (H3N2), HPIV-1, RSV-A*; and iii) other bacteria: *S. mitis, S. pyogenes, S. aureus, E. coli, E. facecaiis*. None of the above interference samples triggered a false positive reaction (Figure 2C). Altogether, these analytical assessments suggest CRISPR-nCoV as a promising molecular assay for 2019-nCoV detection with great sensitivity and specificity.

### Evaluation of CRISPR-nCoV in Clinical Specimens

Upon completion of the analytical assessment, we further evaluated the diagnostic potential of CRIPSR-nCoV in clinical specimens. A total of 114 RNA samples from clinical respiratory samples were included in the evaluation, which consisted of 61 suspected nCoV cases (among which 52 confirmed and 9 ruled-out by mNGS), 17 nCoV-/hCoV+ cases and 36 samples from healthy subjects (Figure 3A).

**Figure 3.**
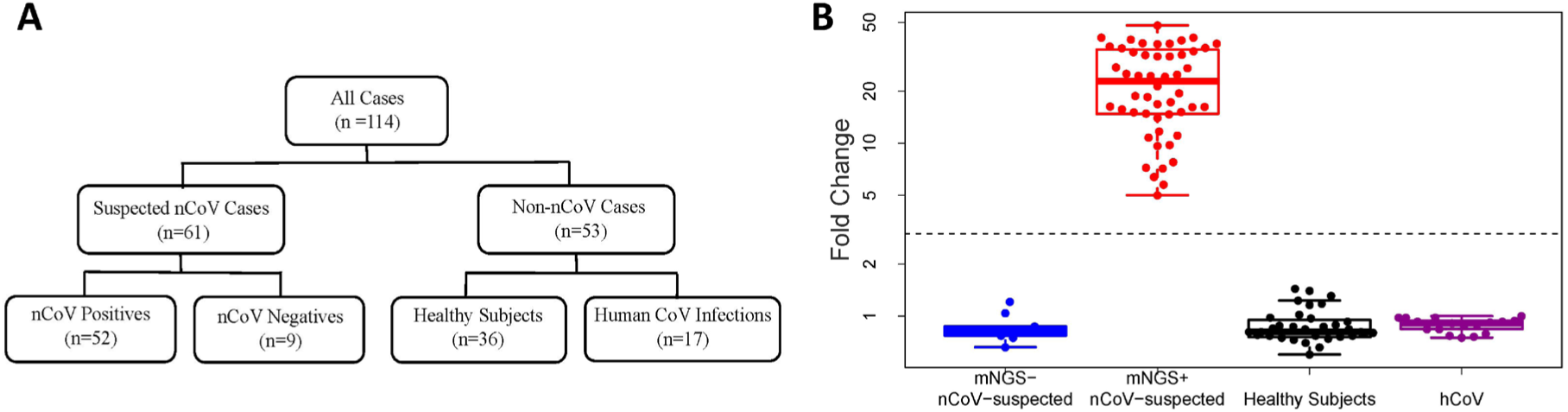
Summary of the cohort and CRISPR-results. (A) A total of 114 specimens included in this study, including 52 cases positive for 2019-nCoV and 62 negative cases. (B) Results of CRISPR-nCoV in different sample groups. Positives and negatives were called based on the fold change and cutoff values.

When conducting the CRISPR-nCoV assay, a positive control (PC) DNA and a no-template control (NC) were included in parallel for each run. Florescent signal from NC was used to normalize the signal and generate corresponding fold-change values (FC). We noticed that there were clear distinction in signal patterns of the reactions. Specifically, the fluorescent signal curve either remained flat as a negative curve (e.g. the NC runs) or had a distinguishable positive signal curve (e.g. the PC runs). The negative curves yielded a maximal FC value of 1.4, whereas the positive ones had a minimal FC value of 5.0. A cut-off was set 3.0 which was set for a complete separation (Figure S1). Consistently, this cut-off offered the optimal sensitivity and specificity as confirmed by an ROC analysis (data not shown).

CRISPR-nCoV demonstrated a sensitivity of 100% by detecting all 52 2019-nCoV-positive cases. No false positives were found in all 62 negative cases, including all the hCoV-infected ones (Figure 3B), suggesting promising clinical sensitivity and specificity of CRISPR-nCoV.

### Diagnostic Performances Among Technological Platforms

We further set out to compare the diagnostic performances among mNGS, RT-PCR and CRISPR. Using mNGS as the reference, PCR- and CRISPR-nCoV both had a specificity of 100% in our study. PCR-nCoV was able to detect the virus in 90.4% (47/52) of the positive cases, with Ct’s ranging from 28.8 to 40.4 and a median Ct of 35.8. It worth noting that the 5 false negative samples by PCR-nCoV had a median mNGS read number of 550, which is much lower than that of the other positive samples at 2,381 reads, suggesting a lower titer of the virus in these samples. CRISPR showed a greater sensitivity by detecting all 52 nCoV-confirmed cases (100%), with FC values ranging from 5.0 to 66.3 and a median FC of 22.8 (Figure 4A).

**Figure 4.**
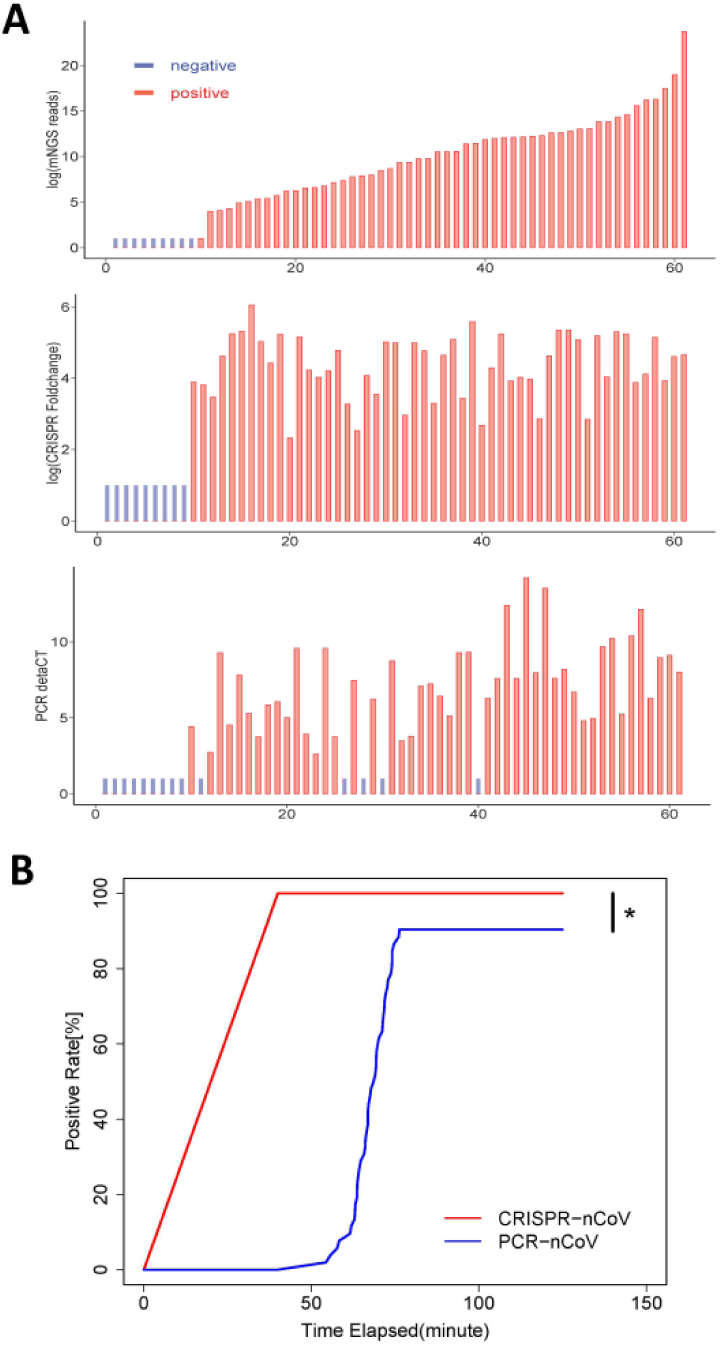
(A) Result summary on the 2019-nCoV-suspected samples by mNGS, CRISPR-nCoV and PCR-nCoV; (B) Kaplan–Meier curve of 2019-nCoV positive rate by CRISPR and PCR. * Log-rank test, *P* < 0.001.

When the reaction turn-around time (TAT) is compared, the CRISPR-nCoV reaction requires only 40 minutes, which is the least among the three and includes 30 minutes of DNA amplification and 10 minutes of Cas reaction. PCR-nCoV requires about 1.5 hours for a completion run of the PCR program. mNGS takes approximately 20 hours, which includes 8 hours of library preparation, 10 hours of sequencing and 2 hours of bioinformatic analysis. Because a positive result may be determined before completing the PCR program, we calculated the effective TAT as the time when the florescent signal reached the threshold. As showed in Figure 4B, CRISPR-nCoV presented a significant advantage in effective TAT over PCR and mNGS.

Altogether, we demonstrated a CRISPR-based assay for 2019-nCoV that offered shorter turn-around time and great diagnostic value, even in under-resourced settings without the need of thermal cyclers. Our study also emphasized strength and weakness of different methodologies which should be fully considered when applied in different diagnostic settings.

## DISCUSSION

Recent progresses in molecular diagnostic technologies, especially mNGS, allowed rapid, initial identification of this novel pathogenic agent at the beginning of the current 2019-nCoV epidemic.[8] Through acquiring the genome sequences of the novel virus, RT-PCR assays were quickly developed for targeted 2019-nCoV detection.[14] However, the sudden outbreak of 2019-nCoV created a dramatic burden not only on the society, but also on public health.

The center of this epidemic, Wuhan city alone, hosts a population of over 10 millions. The surging demand for rapid screening and identification of 2019-nCoV posts a great challenge on the diagnostics.[20] mNGS is the method originally used for the identification of this new viral species and considered as one of the most important references.[8] However, its wider application is limited by its cost and longer TAT of nearly a day. An RT-PCR assay for 2019-nCoV is faster and more affordable. Nevertheless, the need for a thermo cycler by PCR-based diagnostics hinders its use in low-resource settings and curbs the assay throughput. Besides their lower demand for sophisticated temperature controlling instruments, isothermal molecular methods are advantageous owing to its faster nucleic acid amplification. However, there have been debates over the specificity of such methods. The current discovery of the collateral activity of certain Cas family members, provides a great opportunity to take advantage of both the sensitivity of an isothermal assay and the specificity of the CRISPR system.[21] As we demonstrated in this study, CRISPR-nCoV was able to deliver comparable sensitivity and specificity as mNGS within as short as 40 minutes.

As a result of the rapid outbreak, the targeted assays (PCR and CRISPR) were designed and developed based on limited genetic information on 2019-nCoV. Cautions should be taken that certain unknown genomic variations may produce critical impact on the assay efficiency. For example, a mutation or polymorphic site that abrupt the binding of the very 3’ end of a PCR primer may cause a drastic reduction in sensitivity. Although not without its own shortcomings on TAT and costs, NGS-based assays demonstrated a great level of sensitivity and should still be used for continuous monitoring of genetic drifts in the viral genome. These information will provide valuable insights not only from a public health standpoint, but also to guide necessary optimization of targeted assay development.

## Data Availability

data unavailable

